# Intensive care staff perception of medical clowns to an adult intensive care unit: a before-and-after study

**DOI:** 10.64898/2026.10.29.26366802

**Authors:** Daniel J. Jakobson, Hagar Hofesh, Ilya Domanov, Frederic S. Zimmerman

## Abstract

**Introduction:** Medical clowning enhances patient experiences across various clinical contexts. In pediatric ICUs, it reduces stress, sedative use, pain, and family anxiety. It supports pediatric ICU care staff via stress reduction and enhanced workplace morale. Despite this, its application in adult ICUs remains unexplored. This study evaluates staff experiences and perceptions of a novel adult ICU medical clowning program, focusing on staff attitudes before and after implementation and assessing perceived impact on work environment, patients and families.

**Methods:** A medical clown program was initiated in our ICU due to perceived clinical need. The present study took advantage of program initiation to evaluate staff experiences. ICU staff were asked to fill out anonymous questionnaires vis-à-vis their reactions to the program pre- and 6 months post-implementation. Questionnaires were adapted from surveys of clinical staff vis-à-vis medical clowns that were validated in other clinical circumstances. Perceptions of clowning’s effects on patients, families, and staff were assessed using a five-point Likert scale and qualitative feedback.

**Results:** 87 surveys (45 pre-, 42 post-implementation) were assessed. Respondents (74.1% nurses, 27.6% male, mean age 39±2) reported moderately positive attitudes toward clowns (median score 4). Pre-implementation, staff expected moderate benefits for patients, families, and themselves, with stronger effects on patient mood. Post-implementation, perceptions of patient and family effects remained unchanged, but non-significantly improved workplace atmosphere and mood were reported, though differences were not statistically significant. Qualitative feedback highlighted reduced anxiety and calls for program expansion, with some concerns about relevance for unconscious patients.

**Conclusion:** An adult ICU clowning program was well-received, enhancing staff morale without disrupting workflow, suggesting its potential as a non-pharmacological intervention in critical care.

**Key messages:** Medical clowning enhances patient and staff experiences across various clinical contexts, including pediatric ICUs. Despite this, its application in adult ICUs remains unexplored. This study explored the attitude and effects on staff of the initiation of a medical clown program in an adult ICU, showing that program was well-received, enhancing staff morale without disrupting workflow. This suggests its potential as a non-pharmacological intervention in critical care.

## Introduction

Medical clowning was developed in order to improve patient experience in medical situations. In adult patients, medical clowning has been shown to improve lung function in patients with COPD ^1^, increase fertility in patients undergoing in-vitro fertilization ^2^ and help mitigate the impact of several psychological and psychiatric disorders, including post-traumatic stress disorder ^3^ and dementia ^4^, among others ^5^. Medical clowning has been studied in pediatric intensive care units (ICUs), where it may be associated with reduced stress, reduced sedative use and reduced pain. In such units, clowning may also serve to reduce family anxiety and serve as a useful distraction ^6^. Furthermore, physician and nurse burnout in the ICU is a matter concern in multiple studies^7 8^, including limited research in our cultural context^9 10^. Clown interventions may support the work and reduce the stress of the intensive care staff ^11^.

Despite the benefit associated with clowning in both pediatric ICU settings ^6^ and adult non-ICU setting ^11^, to the best of our knowledge medical clowning has not been studied or implemented in the context of an adult ICU. This may be due to perceived interference in the care of these complex and fragile patients, concern regarding nosocomial infection or perceptions regarding the superfluousness of medical clowns in a setting where many patients are sedated ^6^. However, studies of medical staff perceptions of medical clown work in both pediatric and adult settings have generally shown that the medical staff have positive views of the work of medical clowns and their effect on patient care ^5 11 12^.

As part of the initialization of a medical clowns program in our intensive care unit, the staff of our intensive care unit were asked to fill out an anonymous questionnaire about their perceptions of medical clowns in general and in the intensive care unit. This medical clown program and initial questionnaire provide a unique opportunity to study perceptions of intensive care staff on medical clowning in adult intensive care patients and compare these perceptions both before and after incorporation of a medical clown.

Thus, the primary aim of the current study was to assess the attitudes of ICU staff to medical clowning in the intensive care unit and compare attitudes prior to implementation of the ICU medical clowning program versus attitudes after implementation.

## Methods

This prospective, before-and-after single-center survey study, was conducted at the Barzilai University Medical Center (BMC), a 600-bed hospital in Ashkelon, Israel. The study was conducted in the BMC intensive care unit, a 12-bed mixed ICU with both medical and surgical patients. The study was approved by the BMC institutional review board prior to study initiation (approval number: 0026-23-BRZ). Staff surveyed gave their written consent.

Our intensive care unit established a protocol for incorporating medical clowns in the treatment of intensive care patients. In this protocol medical clowns were incorporated into treatment of patients in the intensive care unit and in interactions with their families for 5-15 minutes per a patient per day, during early afternoon hours on weekdays. The medical clowns use circus skills, tricks, and improvisation while making their “rounds” to bring smiles and laughter to patients and their families. These clowns undergo comprehensive training through which they learn both artistic skills and strategies for dealing with psychological issues in the health-care system ^5^. All medically stable patients, as defined by the medical or nursing staff, and all present family members, are eligible for therapy provided by the medical clown. Medical stability included, but was not limited to, hemodynamic stability with or without vasopressors, maintenance of oxygen saturation and sufficient awareness to interact with the medical clown.

In this study we assessed the attitudes and perception of Barzilai University Medical Center Department of Intensive Care staff towards implementation of a medical clowning program in adult ICUs. We compared attitudes and perceptions after implementation of the program versus such attitudes and perceptions that were measured prior to program implementation.

In order to assess such attitudes, the ICU staff were asked to fill out anonymous questionnaires (see supplement) vis-à-vis their reactions to the medical clown program 6 months after initialization of the program. These questions were based on validated surveys of clinical staff vis-à-vis medical clowns in other clinical circumstances ^13^. Staff were surveyed on their impressions of the clown’s effect on patients and on families. They were also surveyed as to the clown’s effect on the respondent. These responses were compared to anonymous questionnaires filled out prior to program initialization, in order to assess and compare attitudes to medical clowning in the ICU.

The data was downloaded to a Microsoft Excel (Ver. 2019) database and then transferred to R (Version 4.4.3. R Foundation for Statistical Computing. released 2024. Vienna, Austria), which was used for analysis.

Comparisons between proportions was performed using the χ^2^-score or the Fisher’s exact test. To compare continuous variables the Student’s t-test or the Mann-Whitney-Wilcoxon test was used. In all tests, two-tailed p-values were taken and a p-value <0.05 was considered significant. A quantitative and qualitative analysis was undertaken of all surveys and comparisons were made between the study survey and the clinical survey undertaken prior to the initiation of the program for all questions.

## Results

49 staff members of the BMC Department of Intensive Care – including physicians, nurses, allied health professionals and nurse aids, but not including researchers – were offered participation in the study. 87 surveys were filled out, 45/49 surveys prior to initiation of the medical clown program and 42/49 surveys 6 months following its initiation. Not all respondents filled out all parts of the survey and some respondents filled out surveys both before and after initiation. 27.6% (24/87) were male, 74.1% (60/81) were nurses and 52.0% (26/50) were employed full time in the BMC ICU. The average age was 39±2 years. Staff reported a moderately positive attitude towards clowns, with a median score of 4 [IQR: 4-4] on a scale of 1 (do not like clowns) to 5 (Very much love clowns). Of responses prior to initiation of the medical clown program 30.2% (13/43) were male and 76.2% (32/42) were nurses. The average age was 36±3. 50.0% (14/28) were full-time in the BMC ICU. A moderately positive attitude towards clowns was reported, with a median score of 4 [3-4]. Of responses 6 months after initiation of the medical clown program 31.4% (11/35) were male and 71.8% (28/39) were nurses. 54.5% (12/22) were employed full-time in the BMC ICU. The average age was 41±4. A moderately positive attitude towards clowns was reported, with a median score of 4 [3-5].A significant difference was noted in age in the survey after implementation versus the survey before (p=0.019), with no other significant differences noted (Table 1).

**Table 1.** Demographics of respondents. CI: confidence interval; IQR: interquartile range.

| Variable | Before | After | p-value |
| --- | --- | --- | --- |
| Male - n (%) | 13/43 (30.2) | 11/35 (31.4) | 1.000 |
| Age (mean ± CI) | 36±3 | 41±4 | 0.019 |
| profession - n (%) | n=42/81 | n=39/81 | 0.857 |
| Nurse | 32 (76.2) | 28 (71.8) |  |
| Physician | 6 (14.3) | 7 (17.9) |  |
| Nurse aid | 1 (2.4) | 2 (5.1) |  |
| Allied health | 3 (7.1) | 2 (5.1) |  |
| Role in the ICU - n (%) | n=28/50 | n=22/50 | 0.618 |
| Full-time | 14 (50.0) | 12 (54.5) |  |
| Part-time | 12 (42.9) | 7 (31.8) |  |
| On call resident | 2 (7.1) | 3 (13.6) |  |
| Personal opinion of clowns – median [IQR] | 4 [3-4] | 4 [3-5] | 0.860 |

In terms of effect on patients, on a five-point scale of 1 (no effect on patients) to 5 (strong effect), prior to program implementation staff expected that medical clowns would have moderate effect in all domains investigated, with a slightly large effect on mood with no differences noted in estimation of effect on patients after program implementation (Figure 1).

**Figure 1.**
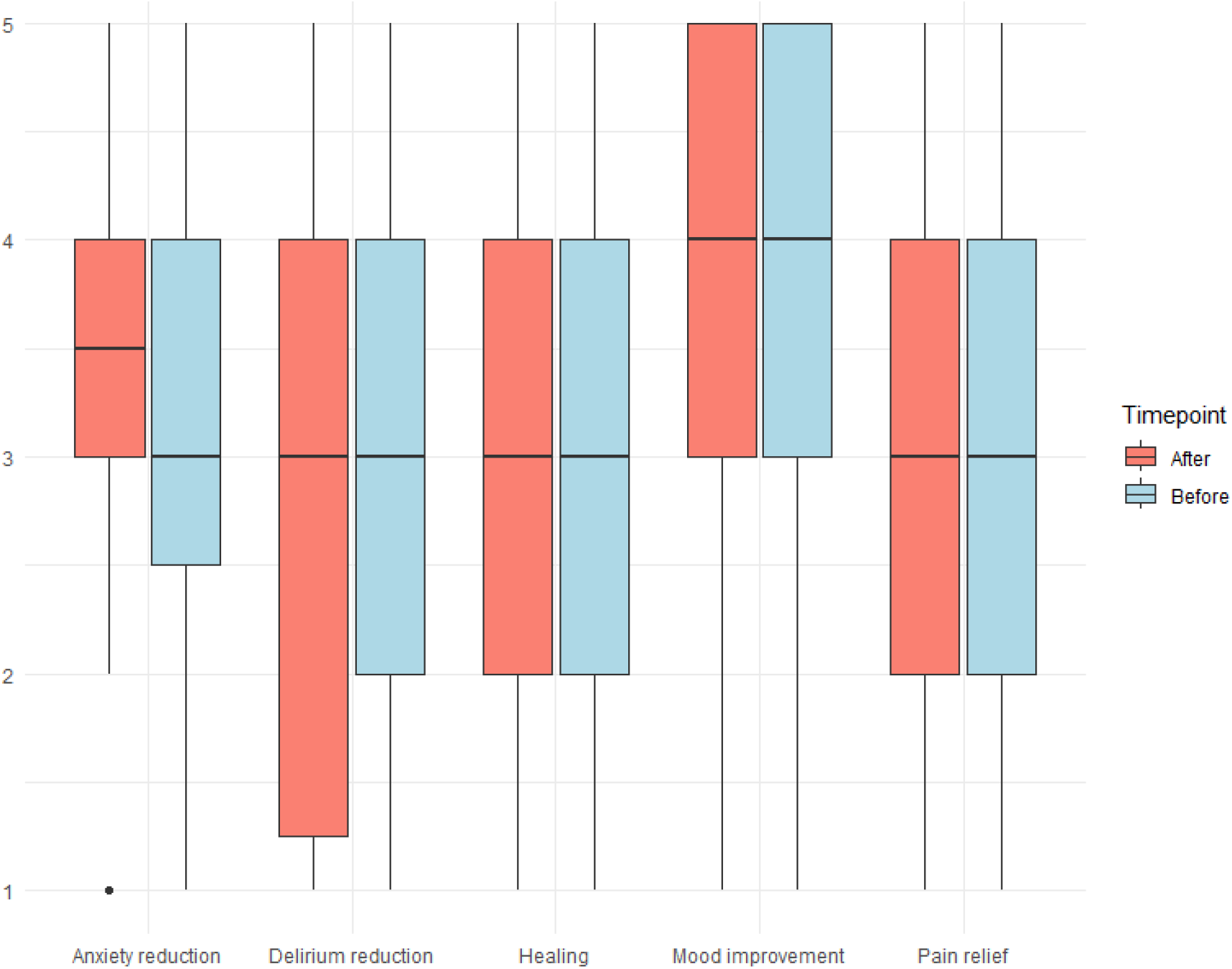
Estimated effect on patients

In terms of effect on families, on a five-point scale of 1 (no effect on families) to 5 (strong effect), prior to program implementation staff expected that medical clowns would have moderate to high effect in positive domains and a moderate to low effect on negative domains. As with patients, no differences noted in estimation of effect on families after program implementation (Figure 2).

**Figure 2.**
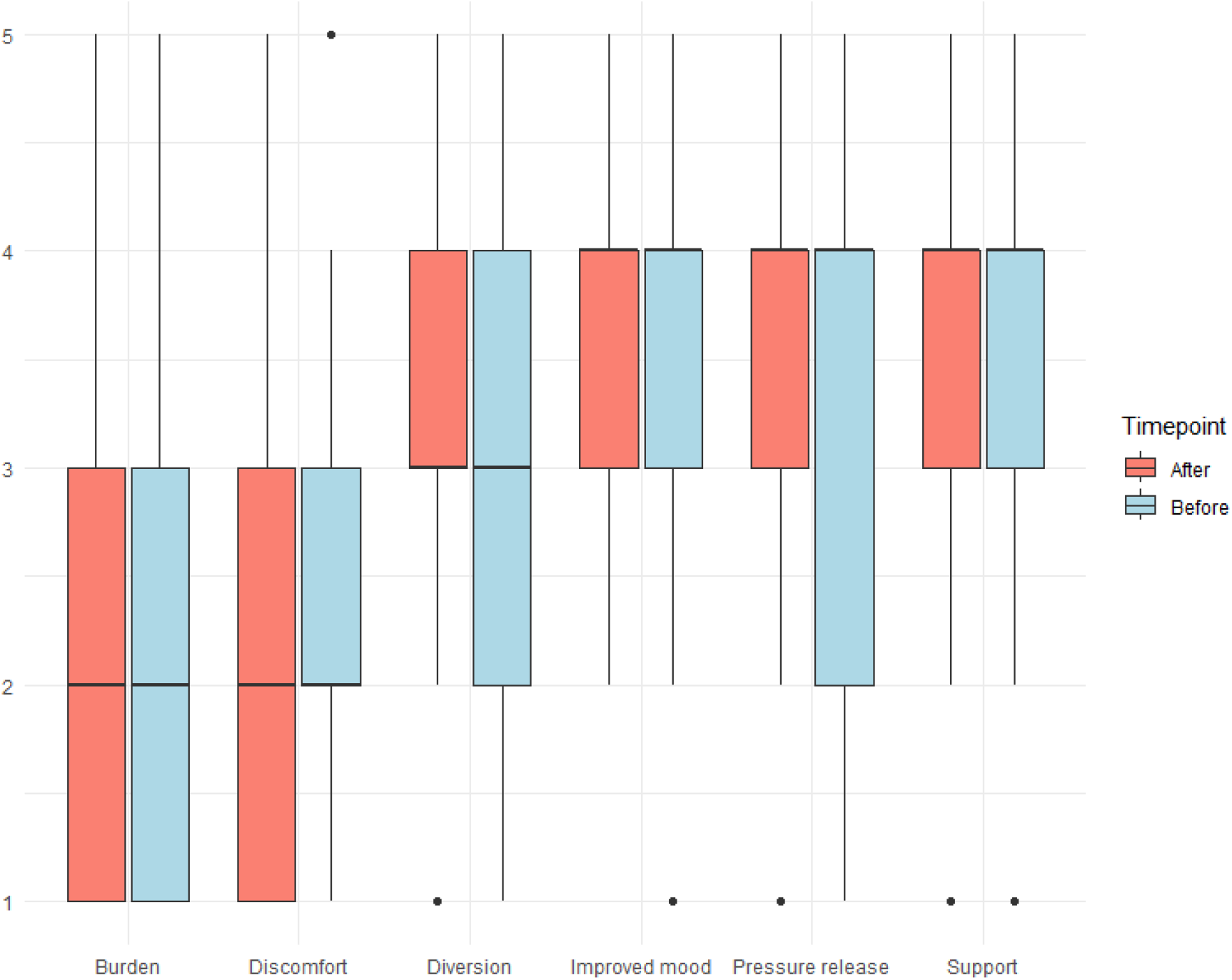
Estimated effect on families

In terms of effect on staff, on a five-point scale of 1 (no effect on staff) to 5 (strong effect), prior to program implementation, staff expected that medical clowns would have moderate effect in positive domains and a moderate to low effect on negative domains. After program implementation, staff reported a moderate to high effect on improved atmosphere and mood and no effect on workflow disruption, though differences did not reach statistical significance as compared to prior to implementation. Effect on pressure release and workload was similar to the effect estimated prior to implementation (Figure 3).

**Figure 3.**
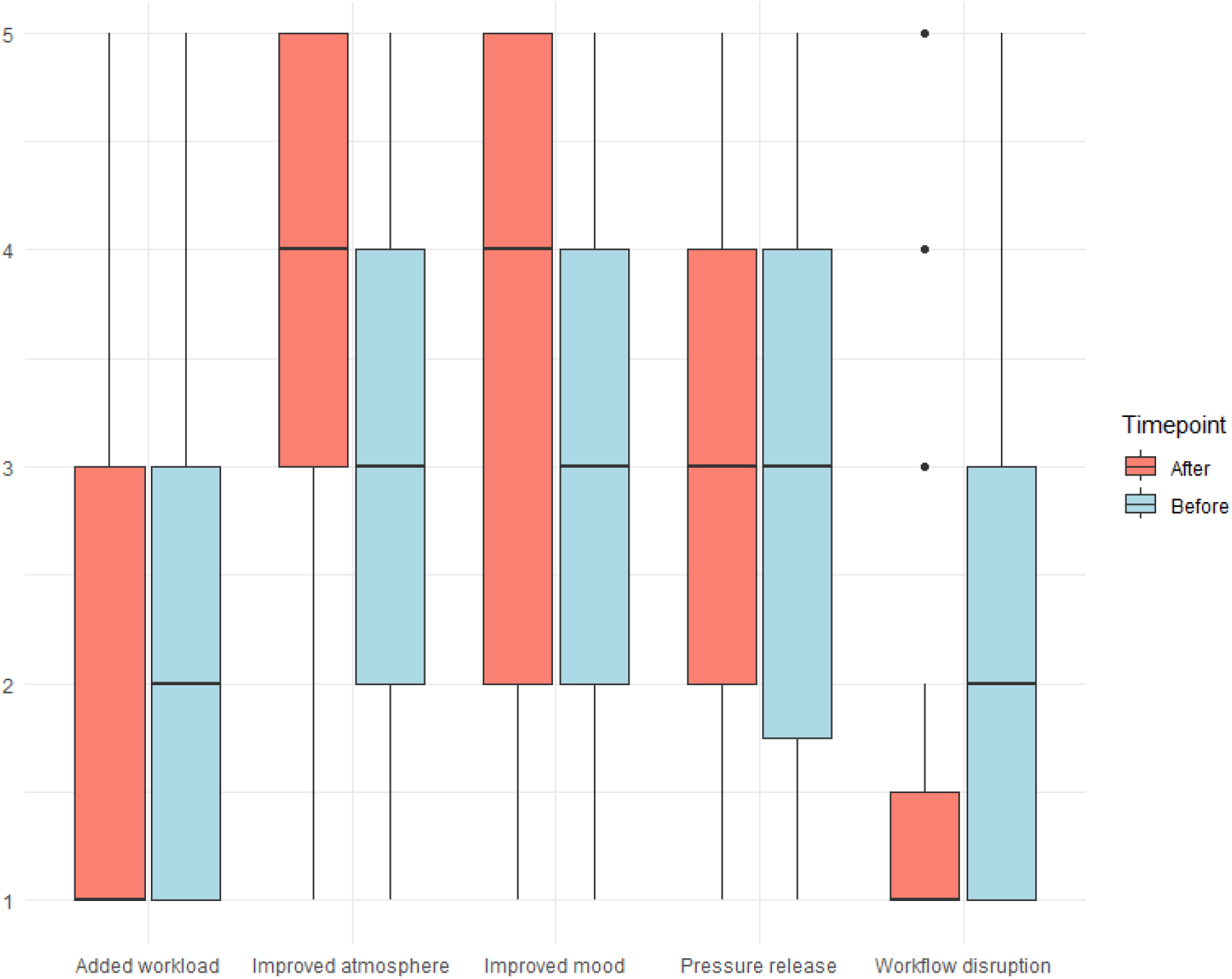
Effect on staff

Qualitative assessments were also sought from participating staff members. Representative positive comments from the survey prior to implementation included: “There is an urgent need for medical clowns in the unit”, “[A medical clown program is] very important especially for younger patients and children, even though I personally do not like clowns”, “[Medical clowns should be included] for patients who have been weaned from mechanical ventilation!”. A representative negative comment was “They do not have to perform while appearing unusual; a regular-looking person with a musical instrument would be more effective.”

Representative positive comments after implementation included: “Clown visits to the department contribute to a pleasant atmosphere for staff, patients and families. Their integration into the department especially with songs, music and laughter alleviates anxiety, reduces stress, and produces a pleasant atmosphere. In my opinion they need to be integrated even more into the regular work of the department” and “Please come every day!!! We love you”. Representative negative comments included “In my opinion it is relevant for children, not for an adult ICU” and “It is not appropriate for every patient. In my opinion, it is only for conscious patients that have already been in the ICU for a couple of days.”

## Discussion

In recent years, survival rates from ICUs have steadily improved ^14 15^ but quality of life after an intensive care stay can often be substantially reduced ^16-19^. It has been postulated that the ICU environment could contribute to the reduction in post ICU quality of life, and, thus, improving the environment and the quality of stay in the intensive care unit could contribute to improved post-ICU outcomes ^14 17 20^. As part of this, medical clowning has been studied in pediatric ICUs, where it may be associated with reduced stress, reduced sedative use and reduced pain as well a tool to reduce family anxiety and as a useful distraction ^6^. In addition, medical clowning can support the work and reduce the stress of the intensive care staff ^11^. Nevertheless, to the best of our knowledge use of medical clowns in an adult ICU has not been studied. Thus, this study was an initial evaluation of staff reaction to implementation of a medical clown program in an adult ICU, both prior to implementation and 6 months after implementation.

In this study, in expectations prior to implementation of the program, staff consistently expected moderate benefits for patients, families, and themselves prior to the program, with a slightly stronger anticipated effect on patient mood. Post-implementation ratings showed no significant change in these estimations for patients and families, suggesting that initial expectations effectively anticipated post-implementation perceptions. However, in terms of effect on staff, a moderate to high effect on improved atmosphere and mood emerged after six months, alongside a reported absence of workflow disruption, both effects that were more positive than anticipated prior to implementation, though these shifts did not reach statistical significance. Nevertheless, this subtle positive impact on the work environment is consistent with findings in other settings, where medical clowns have been shown to enhance staff morale without impeding operational efficiency ^13^. As such, these hypothesis-generating findings should be studied in further research

The quantitative findings of mostly positive effects on patients, staff and family are further enriched by qualitative feedback. Pre-implementation comments revealed a perceived need for clowns, particularly for younger patients or those recovering from critical interventions, despite some personal ambivalence (e.g., “I personally do not like clowns”). Post-implementation, positive remarks emphasized tangible benefits—reduced anxiety, enhanced atmosphere, and calls for greater integration—similar to previous studies highlight clowning’s role in alleviating stress in pediatric and adult healthcare contexts ^13^. Negative feedback, such as the view that clowns are less relevant for adult ICUs or specific patient subsets (e.g., unconscious individuals), aligns with critiques in the literature suggesting that clown interventions may be context- or patient-dependent ^11 21-23^. The suggestion that a “regular-looking person with a musical instrument might be just as effective” reflects a preference for less theatrical interventions among some staff, potentially indicating a need for customizable clowning approaches.

The implications of these findings are twofold. First, the sustained positive attitude and reported atmospheric improvements support the integration of medical clown programs as a low-disruption, possibly morale-boosting intervention in adult ICUs, extending their application beyond their traditional pediatric and non-critical care settings. Secondly, the lack of significant change in perceived patient and family outcomes was noted post-implementation. This may suggest that staff (perhaps especially nursing staff from whom the majority of feedback was received) may be effective arbitrators of the potential usefulness of non-pharmacological inventions such as medical clowning. Alternatively, perhaps an intervention spanning more time or utilizing more direct evidence could demonstrate broader impacts.

Overall, this study contributes to the growing body of evidence on medical clowning by demonstrating its feasibility in a critical care context, with nuanced staff-specific and possible family and patient benefits.

Several limitations must be considered. The modest sample size (n=87 total surveys) and incomplete responses reduce the statistical power to detect subtle differences, particularly in effect ratings and may also introduce a sampling bias. Additionally, the overlap of respondents completing both pre- and post-implementation surveys introduces potential bias, as their prior opinions may have influenced later responses. Staff understandings of patient perceptions might not accurately reflect patient experiences, and the absence of patient or family perspectives further limits the ability to triangulate staff perceptions with direct outcomes. Further studies are planned to more directly assess patient and family experiences. Finally, the six-month timeframe may be insufficient to capture long-term effects on staff attitudes or patient care metrics, such as length of stay or recovery rates. Future research could address these gaps by employing larger, longitudinal designs that include diverse respondent groups (e.g., patients, families, and staff) and objective measures such as agitation and pain scale scores, as well as use of analgesics and sedatives. We hope to address some of these issues in further research in our institution.

In conclusion, this study demonstrates that a medical clown program in the BMC ICU was well-received by staff, with indications of enhanced workplace atmosphere and mood six months after implementation and without workflow disruption, with possible broader effects on patients and families. These findings underscore the program’s potential as a supportive tool in critical care and as possible non-pharmacological means of enhancing ICU outcomes.

## Data Availability

All data produced in the present study are available upon reasonable request to the authors

